# Clinical Manifestations of Children with COVID-19: a Systematic Review

**DOI:** 10.1101/2020.04.01.20049833

**Authors:** Tiago H. de Souza, José A. Nadal, Roberto J. N. Nogueira, Ricardo M. Pereira, Marcelo B. Brandão

## Abstract

**Background:** The coronavirus disease 2019 (COVID-19) outbreak is an unprecedented global public health challenge, leading to thousands of deaths every day worldwide. Despite the epidemiological importance, clinical patterns of children with COVID-19 remain unclear. The aim of this study was to describe the clinical, laboratorial and radiological characteristics of children with COVID-19.

**Methods:** The Medline database was searched between December 1^st^ 2019 and April 6^th^ 2020. No language restrictions were applied. Inclusion criteria were: (1) studied patients younger than 18 years old; (2) presented original data from cases of COVID-19 confirmed by reverse-transcription polymerase chain reaction; and (3) contained descriptions of clinical manifestations, laboratory tests or radiological examinations.

**Results:** A total of 38 studies (1,124 cases) were included. From all the cases, 1,117 had their severity classified: 14.2% were asymptomatic, 36.3% were mild, 46.0% were moderate, 2.1% were severe and 1.2% were critical. The most prevalent symptom was fever (47.5%), followed by cough (41.5%), nasal symptoms (11.2%), diarrhea (8.1%) and nausea/vomiting (7.1%). One hundred forty-five (36.9%) children were diagnosed with pneumonia and 43 (10.9%) upper airway infections were reported. Reduced lymphocyte count were reported in 12.9% of cases. Abnormalities on computed tomography was reported in 63.0% of cases. The most prevalent abnormalities reported were ground glass opacities, patchy shadows and consolidations. Only one death was reported.

**Conclusions:** Clinical manifestations of children with COVID-19 differ widely from adults cases. Fever and respiratory symptoms should not be considered a hallmark of COVID-19 in children.

## INTRODUCTION

In late December 2019, Chinese authorities informed the World Health Organization (WHO) that, due to unknown cause, an outbreak of pneumonia emerged in Wuhan, Hubei province. On January 7, 2020, a new type of coronavirus (severe acute respiratory syndrome coronavirus 2, SARS-CoV-2) was isolated and few days after the disease has been named “coronavirus disease 2019” (abbreviated “COVID-19”). The first death caused by COVID-19 was on January 9, 2020, in Wuhan and since then more than 370,000 cases and 16,000 deaths occurred worldwide.^1^ Nowadays, the death toll in Italy has exceeded four timese the number registered in China and the United States became the new COVID-19 epicenter.

In spite of stepped-up efforts to contain the pandemic, the number of affected patients and the death toll continue to rise. Elderly patients infected with SARS-CoV-2 are at high risk to have severe acute respiratory syndrome, complications and death.^2^ Due to unknown reasons, children with COVID-19 appear to have a milder clinical course compared to adults, and reports of death are scarce.^3,4^ However, pediatric population may play a major role in community spread of SARS-CoV-2. In addition to viral shedding in nasal secretions, there is evidence of fecal shedding for several weeks after diagnosis, which poses a challenge for infection control.^5^

Despite the epidemiological importance, clinical patterns of children with COVID-19 remain unclear. The WHO recommends testing all suspected cases, however, children infected with SARS-CoV-2 may not meet all the criteria required in the suspected case definition.^6^ The objective of this study is to describe the clinical, laboratorial and radiological characteristics of children with COVID-19 reported in the literature.

## METHODS

This review was performed in accordance with the Preferred Reporting Items for Systematic and Meta-Analysis (PRISMA) statement.^7,8^ The Medline database was searched using the following search strategy: ((((covid-19) OR coronavirus) OR SARS-CoV-2)) AND (((((((pediatrics) OR children) OR neonates) OR child) OR neonate) OR infant) OR infants). No language restrictions were applied. Articles published between December 1^st^ 2019 and April 7^th^ 2020 were evaluated for inclusion. No attempts were made to contact the study authors for identifying missing and confusing data. A manual search of the references found in the selected articles and reviews was also performed.

### Study Selection

Two authors (THS and JAN) screened the titles and abstracts independently and in duplicate for potential eligibility. They subsequently read the full texts to determine final eligibility. Discrepancies were resolved through discussion and consensus, and if necessary, the assistance of a third author (MBB) was sought.

Eligible studies fulfilled the following criteria: (1) studied patients younger than 18 years old; (2) presented original data from cases of COVID-19 confirmed by reverse-transcription polymerase chain reaction; (3) contained descriptions of clinical manifestations, laboratory tests or radiological examinations.

### Data Extraction

A structured data extraction form was piloted and then used to extract data from the reports of all included studies in duplicate and independently by two authors (THS and JAN). Discrepancies in extracted data were resolved through discussion. The following data were extracted, when available, from each elected article: first author, publication year, study design, number of cases, gender, age, clinical manifestations, laboratory tests, radiological examinations and outcomes (discharged, still hospitalized or death).

When sufficient data was reported, the cases were classified into the following clinical types:^9^

1. Asymptomatic infection: without any clinical symptoms and signs and the chest imaging is normal, while the SARS-CoV-2 nucleic acid test was positive or the serum-specific antibody was retrospectively diagnosed as infection.
2. Mild: symptoms of acute upper respiratory tract infection, including fever, fatigue, myalgia, cough, sore throat, runny nose, and sneezing. Physical examination shows congestion of the pharynx and no auscultatory abnormalities. Some cases may have no fever, or have only digestive symptoms such as nausea, vomiting, abdominal pain and diarrhea.
3. Moderate: presented as pneumonia. Frequent fever and cough, mostly dry cough, followed by productive cough, some may have wheezing, but no obvious hypoxemia such as shortness of breath, and lungs can hear sputum or dry snoring and / or wet snoring. Some cases may have no clinical signs and symptoms, but chest computed tomography (CT) shows lung lesions, which are subclinical.
4. Severe: Early respiratory symptoms such as fever and cough, may be accompanied by gastrointestinal symptoms such as diarrhea. The disease usually progresses around 1 week, and dyspnea occurs, with central cyanosis. Oxygen saturation is less than 92%, with other hypoxia manifestations.
5. Critical: Children can quickly progress to acute respiratory distress syndrome (ARDS) or respiratory failure, and may also have shock, encephalopathy, myocardial injury or heart failure, coagulation dysfunction, and acute kidney injury, including multiple organ dysfunction. Can be life threatening.

## RESULTS

### Study Selection and Characteristics

Of 293 potentially relevant articles identified by the search strategy, 38 met the inclusion criteria. A total of 1117 descriptions of pediatric cases^3,5,18–27,10,28–37,11,38–40,12– 17^ and 7 neonate cases^41–45^ of COVID-19 were obtained, being that 643 were males 478 were females, and 3 were not disclosed. The flow diagram (**Figure 1**) summarizes the steps followed to identify the studies meeting the inclusion criteria.

**Figure 1.**
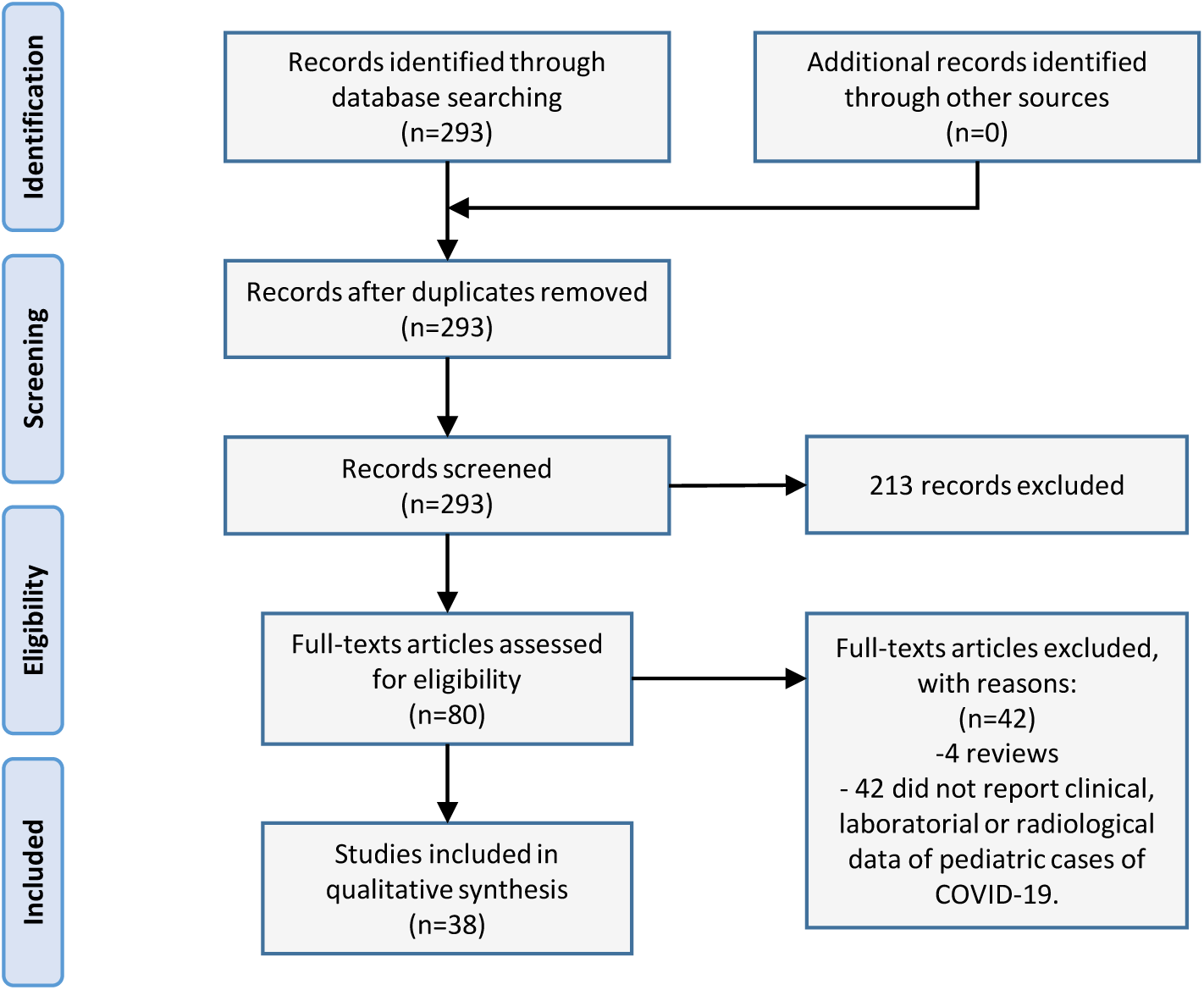
Preferred Reporting Items for Systematic Reviews and Meta-Analyses flow diagram of selection process.

Twenty studies were case reports,^20,21,32,35,37–43,45,22–28,31^ 11 were case series^5,15,44,16–19,29,30,33,34^ and 6 were retrospective studies.^3,10–14,36^ With the exception of 5 multicenter studies,^5,10–12,36^ all others were unicentric studies. Thirty-three studies were conducted in China,^3,5,18–22,24–27,29,10,30,32–37,39–41,11,42–44,12–17^, 1 in Italy,38 1 in Iran,^45^ 1 in Singapore,^28^ 1 in Korea,^23^ and 1 in Vietnam.^31^

### Severity of illness

A total of 1117 cases had their severity classified based on the reported clinical data. One hundred fifty-nine (14.2%) cases were asymptomatic, 406 (36.3%) were mild, 514 (46.0%) were moderate, 25 (2.1%) were severe and 13 (1.2%) were critical cases. **Table 1** summarizes the severity of illness reported in each included study.

**Table 1.**
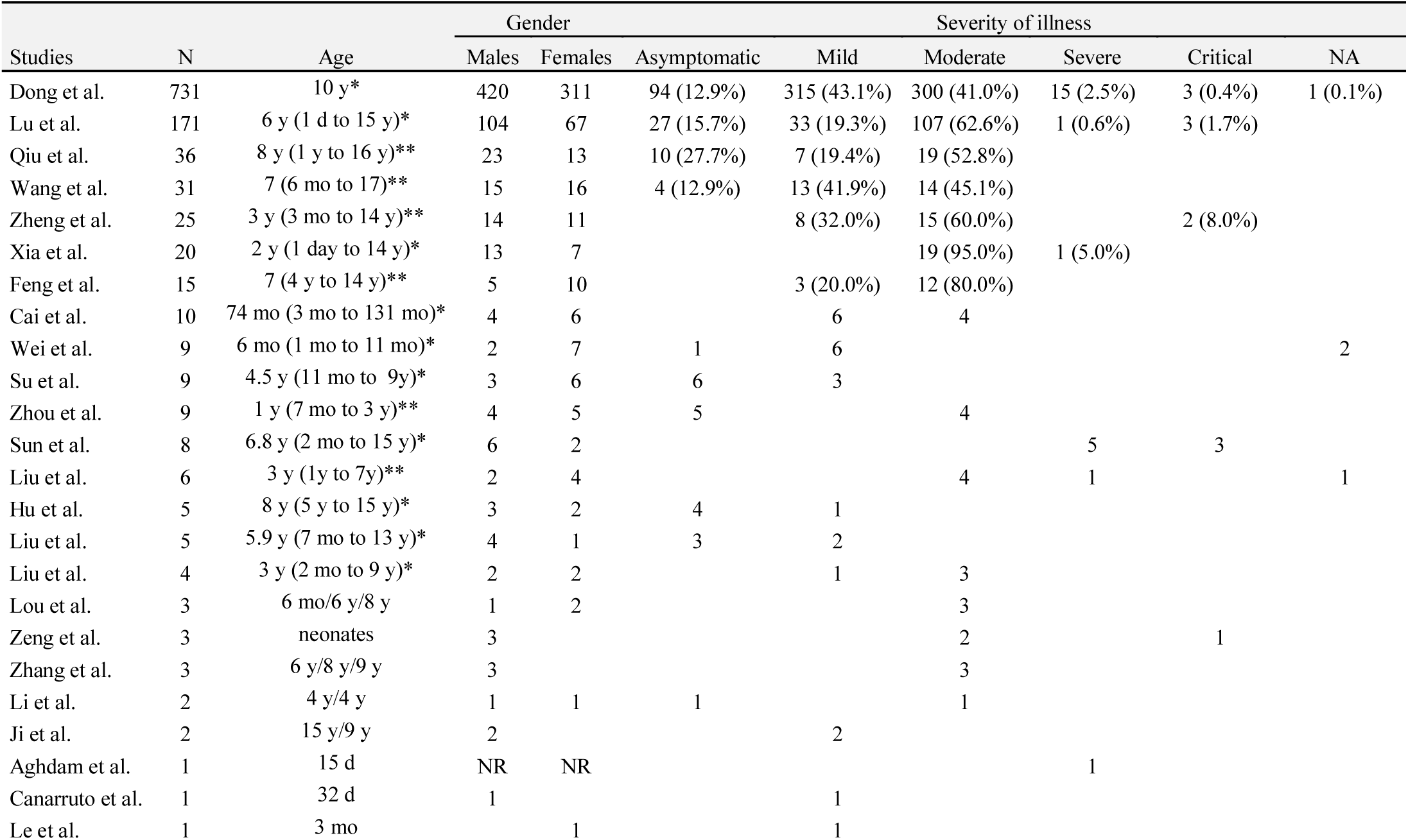
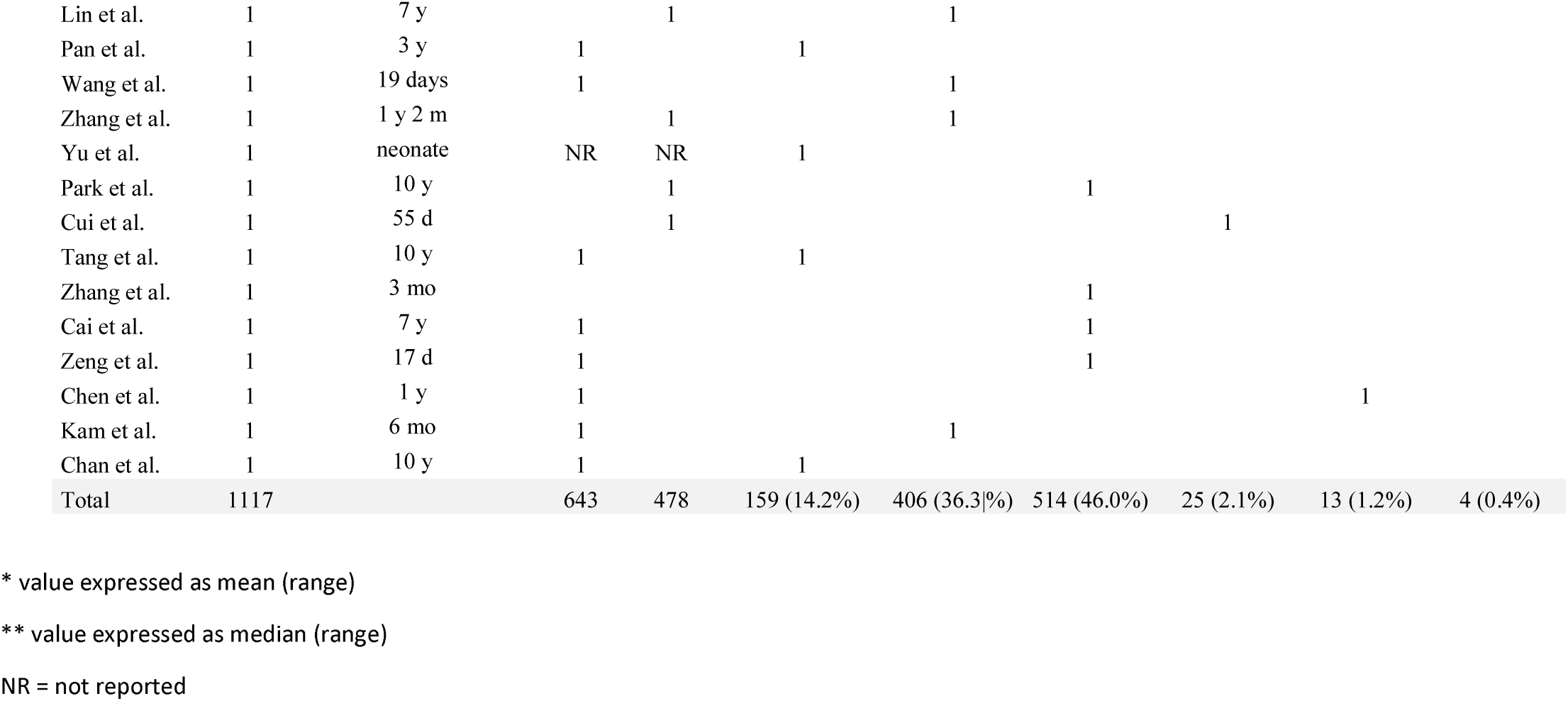
Severity of illness and characteristics of cases reported.

### Clinical manifestations

The most prevalent symptom was fever, reported in 47.5% of the cases, followed by cough (41.5%), nasal symptoms (11.2%), diarrhea (8.1%), nausea/vomiting (7.1%), fatigue (5.0%) and respiratory distress (3.5%). One hundred forty-five (36.9%) children were diagnosed with pneumonia and 43 (10.9%) upper airway infections were reported. Amongst the most common clinical signs described were pharyngeal erythema (20.6%), tachycardia (18.6%) and tachypnea (13.4%) on admission. All the clinical manifestations reported in the selected studies and their relative frequencies are described in **Table 2**. All clinical manifestations described in each study are presented in **E-Table 1** in the Supplement.

**Table 2.**
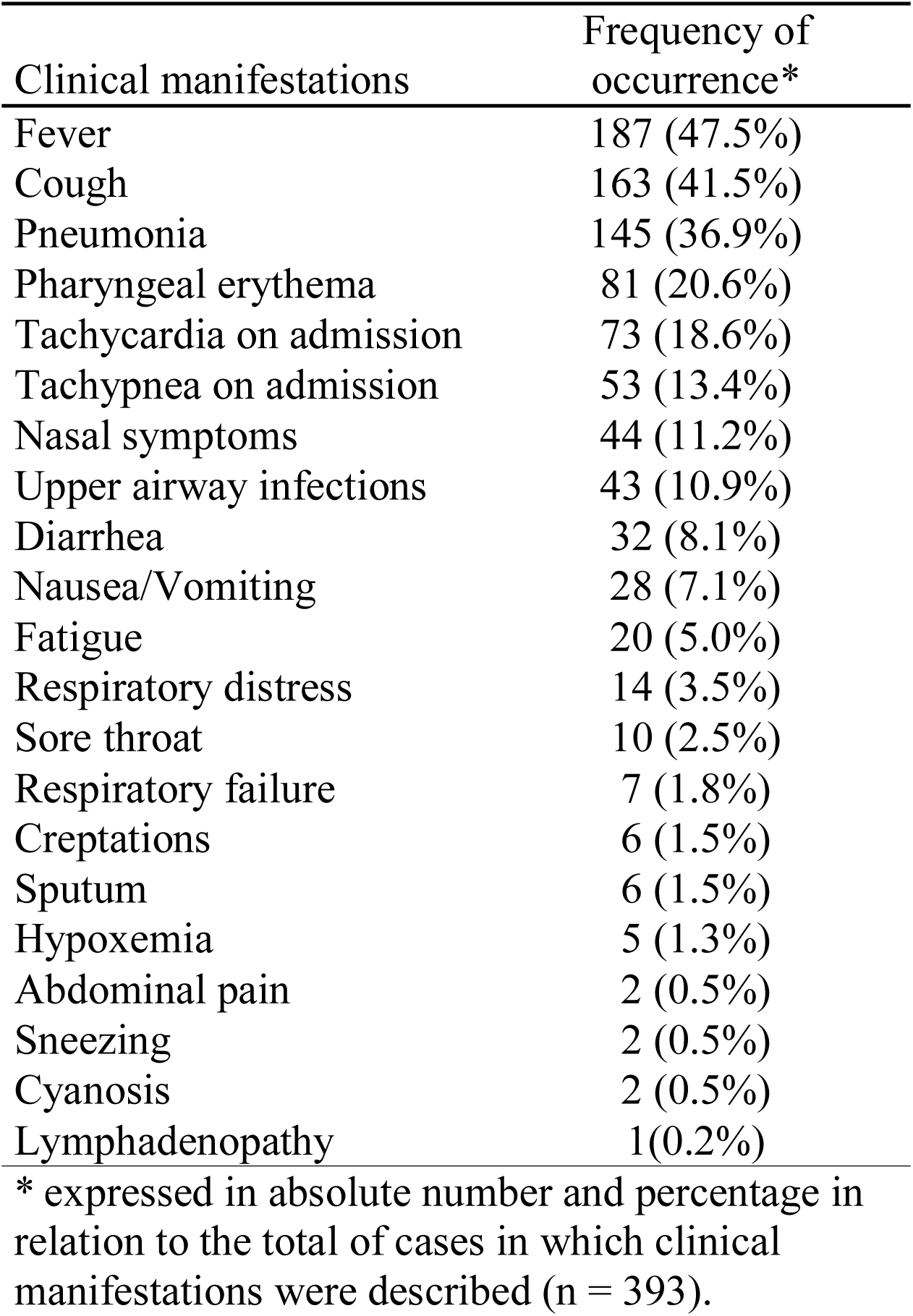
Distributions of clinical manifestations of children with COVID-19 described in the selected studies.

### Laboratorial characteristics

Twenty-nine studies detailed the white blood cell count of 174 cases.^11,13,24– 33,14,34–43,16,45,17–19,21–23^ Of these, 13 (7.5%) were elevated, 29 (16.6%) were decreased, and 132 (75.8%) were within normal ranges. Normal or elevated lymphocytes count were detailed in 28 studies, involving 154 cases.^11,13,26–35,16,36–43,45,17,19,21–25^ Normal lymphocyte count were reported in 69.5% and elevated in 11.7% of cases. Reduced lymphocyte count were reported in 45 of 350 (12.9%) cases.^3,11,24–33,12,34–43,13,45,16,17,19,21–23^. Zheng et al reported a median white blood cell count of 6.2 × 10^9^/L [interquartile range (IQR) 4.30–9.85] and median lymphocyte count of 2.19 × 10^9^/L (IQR 1.15–3.31) of 25 pediatric patients.

Normal or decreased platelet count were detailed in 15 studies, involving 32 cases.^16,23,37,39–41,45,24,25,27,29,31–33,35^ Normal platelet count were reported in 78.1% (22/32) and descreased in 3.2% (1/32) of cases. Increased platelet count were reported in 6 of 63 (9.5%) cases.^11,16,35,37,39–41,45,23–25,27,29,31–33^

*Elevated C-reactive protein (CRP) levels were described in 59 of 305 cases* (19.3%), reported in 25 studies.^3,11,26,27,29,31–37,13,39–42,45,16,19–24^. Zheng et al reported a median CRP level of 15.5 mg/dL (IQR 0.93-25.04) in 25 cases, and Cai et al. a median of 7.5 mg/dL in 10 cases.

Procalcitonin levels were elevated in 139 of 279 cases (49.8%), reported in 16 studies.^3,11,37,39–43,13,16,24,26,27,31,33,36^ Cai et al reported a median procalcitonin level of 0.07 ng/dL in 10 cases.

Increased liver enzymes were described in 56 of 292 cases (19.2%), reported in 16 studies.^3,11,34–37,39–41,43,13,16,21,24,27,29,31,33^ Zheng et al reported a median of 12 U/L in 25 cases, and Cai et al. a median alanine aminotransferase and aspartate aminotransferase level of 18.5 U/L and 27.7 U/L, respectively, in 10 cases.

Co-infections with other pathogens were reported in 5 studies.^12,13,16,19,34^ Two patients were reported with influenza A, 5 with influenza B, 3 with respiratory syncytial virus (RSV), 1 with cytomegalovirus, 7 with *Mycoplasma pneumoniae* and 1 with *Enterobacter aerogenes*.

### Radiological features

Twenty-seven studies reported 184 cases which underwent chest CT.^3,11,21– 27,29,30,32,12,34–36,39–42,13,14,16–20^ One hundred sixteen (63.0%) CT scans presented abnormalities. The most prevalent abnormalities reported were ground glass opacities, patchy shadows and consolidations. In the study of Lu et al. involving 171 cases, ground glass opacities and patchy shadowings were observed in 32.7% and 31% of cases, respectively.^3^ Pleural effusion was observed in a 2-month-old child with simultaneous RSV and SARS-CoV-2 infections.^19^

### Outcomes

Clinical outcomes of death, discharged or still hospitalized were described for 371 cases in 32 studies.3,5,21–29,31,11,33–42,12,44,45,13,14,16–18,20 Of these, 62 cases were still hospitalized when studies were submitted, 308 were discharged and 1 died.

## DISCUSSION

In our study, we described the main clinical, laboratorial and radiological characteristics of children infected with SARS-CoV-2 reported in the literature. It was observed that only a small proportion of infected children became severely or critically ill. About half of the children with COVID-19 were asymptomatic or mild cases, and several were classified as moderate due to radiological abnormalities in spite of their mild clinical manifestations. The prognosis seems to be very good, with recovery described in the vast majority of reported cases. Only one death was reported in the included studies, a 10-month-old child with intussusception.^3^

Since COVID-19 has a favorable clinical course in children, the importance of pediatric cases is mainly due to epidemiological issues. Despite being mild or asymptomatic cases, prolonged viral shedding in stool and nasal secretions made children facilitators of viral transmission.^5,46^ In the study of Xu et al., eight of ten children with SARS-CoV02 had persistently positive rectal swabs even after their nasopharyngeal tests were negative.^46^ This raises concerns about the possibility of a fecal–oral route of transmission. The role of children in the transmission chain needs to be urgently clarified to establish social and public health policies for the protection of vulnerable populations, such as the elderly and people with comorbidities.

Testing people who meet the COVID-19 suspected case definition is essential for clinical management and outbreak control. The Centers for Disease Control and Prevention (CDC) recommends that clinicians should decide to test patients based on the presence of signs and symptoms compatible with COVID-19. The WHO, CDC and several other government health agencies emphasize fever and respiratory symptoms in the criteria for suspected cases, however, we observed in our study that only 47.5% of pediatric cases had fever.^47,48^ Since many are asymptomatic or mild cases, children certainly are not tested as often as adults, leading to an underestimate of the true numbers of infected people and increased transmission of the virus.

Guan et al. demonstrated pronounced lymphopenia in adults with COVID-19, especially in severe cases, where the observed prevalence was 96.1%.^4^ Some authors even suggest that lymphopenia is a predictor of prognosis in adult patients with COVID-19.^49^t In our study, decreased lymphocyte count was described in only 12.9% of infected children, in contrast with adults, in which 80% of the non-severe cases have lymphopenia. With this, lymphopenia may not be a reliable indicator of COVID-19 in children.

Similarly to adults, the most prevalent abnormalities on chest CT of children with COVID-19 were ground glass opacities and patchy shadowings. However, while 86.2% of adults cases presented any abnormalities on chest CT, the same occurred in only 63.0% of children in the selected studies. Descriptions of chest x-rays of pediatric cases are scarce and would be useful for resource-limited settings.

Our study has some limitations. Firstly, data from the same patient may have been presented in more than one included study. Secondly, the majority of data are from China, and may not be generalized for other populations.

## CONCLUSION

Most children with COVID-19 have a favorable clinical course and their clinical manifestations differ widely from adults cases. Fever and respiratory symptoms should not be considered a hallmark of COVID-19 in children. With this, pediatricians should have a high level of clinical suspicion to diagnose children infected with SARS-Cov-2, as the majority of pediatric cases are asymptomatic or mild. Regardless of the favorable prognosis, it is important that the child’s role in the contamination chain is precisely established and considered.

## Data Availability

All relevant data are described in the manuscript.

## Abbreviations

WHO: World Health Organization
SARS-CoV-2: Severe acute respiratory syndrome coronavirus 2
COVID-19: Coronavirus disease 2019
CT: Computed tomography
CDC: Centers for Disease Control and Prevention

## Acknowledgements

Thank you to Carolina Grotta Ramos Telio for her review of the manuscript.

---

**E-Table 1.**
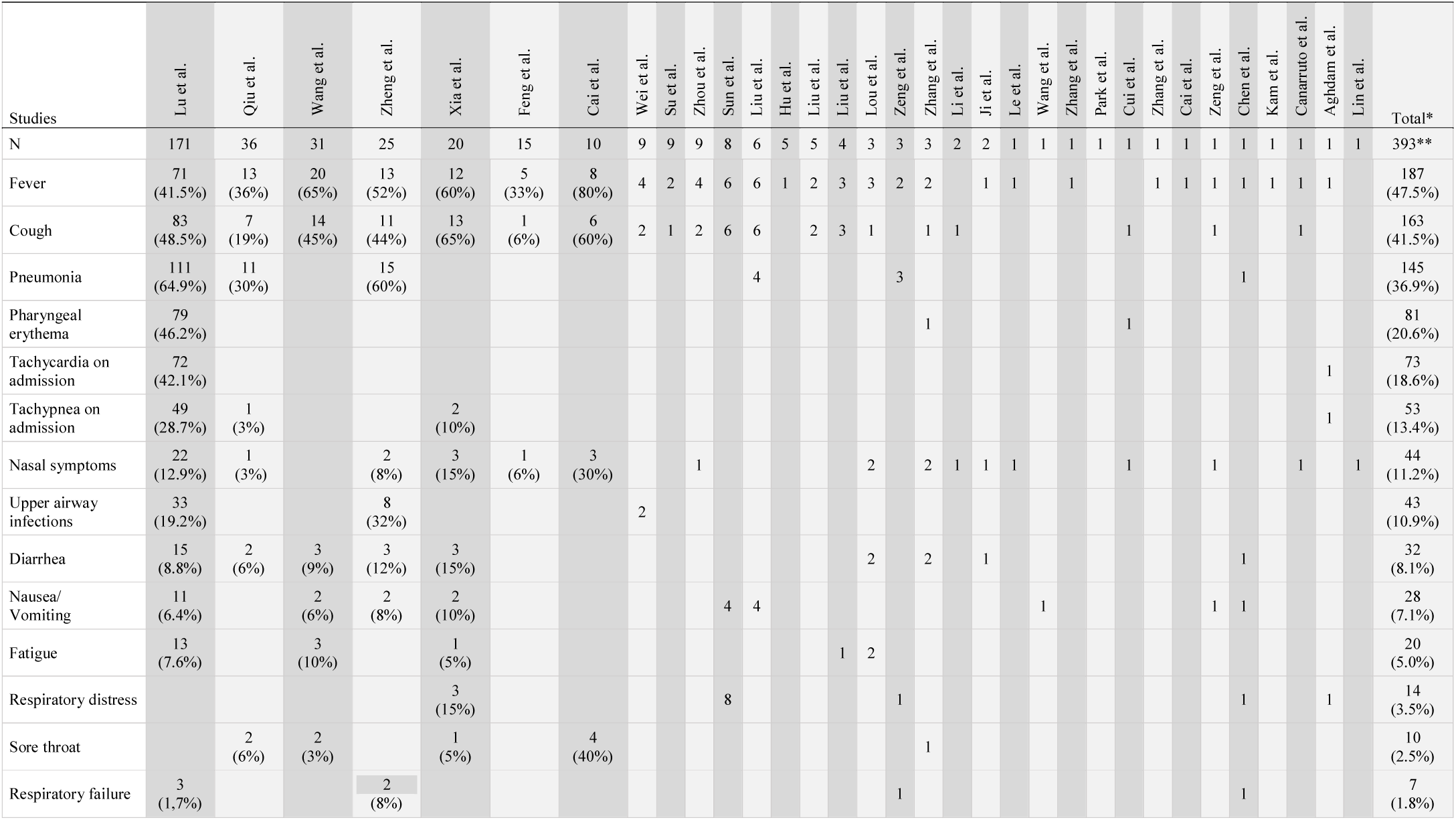
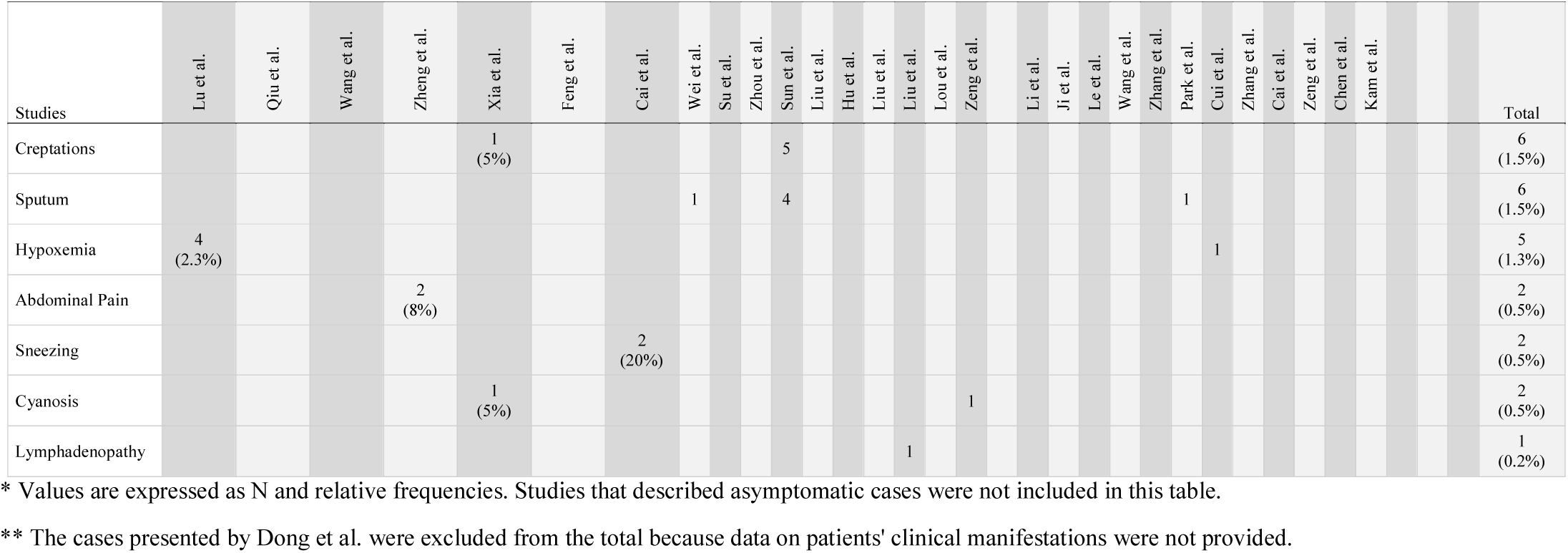
Clinical manifestations reported in the selected studies.

